# Methicillin-Susceptible *Staphylococcus aureus* ST398 in atopic dermatitis in Portugal displays pathogenic traits associated with impaired skin barrier function

**DOI:** 10.64898/2026.02.17.26346495

**Authors:** Diana Caieiro, Nuno A. Faria, Ana Botelho, Mariana Araújo, Leonor Ramos, Joana Calvão, Margarida Gonçalo, Maria Miragaia

## Abstract

*Staphylococcus aureus* plays a central role in the exacerbation of atopic dermatitis (AD), but the population structure and pathogenic determinants of strains colonizing AD patients remain poorly understood. It is unclear whether these strains mirror those circulating in the general community or whether specific clonal lineages are selectively adapted to the AD skin microenvironment. Data addressing this question are scarce, particularly in Portugal. In this study, we investigated the molecular epidemiology and pathogenic traits of *S. aureus* colonizing skin lesions in adult patients with AD in Portugal. We found that lesion-associated isolates belonged predominantly to the methicillin-susceptible *S. aureus* MSSA-ST398 clonal type, a lineage that is widely circulating in the Portuguese community, particularly among vulnerable populations, and that has also been implicated in severe human infections. Notably, isolates from this clonal type in AD harboured specific pathogenicity traits associated with skin barrier disruption, including hemolysin and urease production, which may contribute to their success as colonizers in AD. Our findings highlight that *S. aureus* colonization in AD arises from a dynamic interplay between community-level molecular epidemiology and disease-specific selective pressures. While circulating lineages provide the genetic background diversity, the AD skin microenvironment appears to shape which clones ultimately become dominant. Such an integrated perspective may help to inform future geographically tailored strategies aimed at limiting bacterial burden and preventing disease exacerbation in AD.

## To the Editor

*Staphylococcus aureus* plays a major role in atopic dermatitis (AD) exacerbation, but the underlying mechanisms remain incompletely understood (Chehadeh et al. 2026). It is unclear whether *S. aureus* strains colonizing AD patients merely reflect those circulating as colonizers of healthy humans in the community or whether specific clonal lineages are better adapted to the microenvironment of AD skin. The population structure of *S. aureus* in AD remains poorly characterized, including in Portugal. Addressing this gap may inform targeted local strategies to reduce consequent disease exacerbation. We therefore provide the first detailed characterization of *S. aureus* colonization, clonal lineages, antibiotic resistance, and virulence traits in Portuguese AD patients.

This study was approved by the Ethics Committee of the Hospital. Written informed consent was obtained from all participants. Between April 2024 and April 2025, adult AD patients (>18 years), diagnosed according to Hannifin and Rajka criteria, were recruited during routine visits to the AD consultation of the Dermatology Department at Coimbra University Hospital. Saline-prewetted cotton swabs were used to sample the nares (~1 cm^2^) and 2 cm^2^ of lesional and non-lesional skin per patient. *S. aureus* was isolated using a culture-based assay (Supplementary Methods).

Fifty-seven AD patients (28 males/29 females; median age, 30 years) were enrolled. Eczema Area and Severity Index (EASI) scores ranged from 0.5 to 50 (median, 10) and Worst Itch Numerical Rating Scale (WI-NRS) from 0 to 10 (median, 5.5). No skin lesion showed clinical signs of secondary infection at sampling. Dupilumab was received by 43.9% (25/57) of patients, and conventional systemic therapy by 33.3% (19/57).

Most AD patients (82%; 47/57) were colonized by *S. aureus* in at least one sampling site (Figure 1a), most commonly involving lesional skin, either with nasal carriage (14/57) or with both nasal and non-lesional skin colonization (12/57). Nasal carriage without detectable skin colonization occurred in 11 patients, all with EASI scores <7 and receiving dupilumab or conventional systemic therapy. Overall, all colonization patterns across the three sites were observed, highlighting the heterogeneity of *S. aureus* colonization in AD. By site, carriage was most frequent in the anterior nares (74%; 42/57), followed by lesional (53%; 30/57) and non-lesional skin (21%; 19/57), consistent with previous reports of high lesional skin colonization (Totté et al. 2016). However, colonization of non-lesional skin in our study was substantially higher than the rate described for healthy individuals, in whom *S. aureus* is typically absent (Piewngam and Otto 2024). Although the role of the anterior nares as a reservoir of *S. aureus* in AD patients has been previously suggested (van Mierlo et al. 2021), our whole genome based single nucleotide polymorphism (SNPs) analysis showed that isolates from nares and lesions within the same patient were highly genetically related (<16 SNPs), further supporting the nares as a putative source (supplementary Table S2).

**Figure 1.**
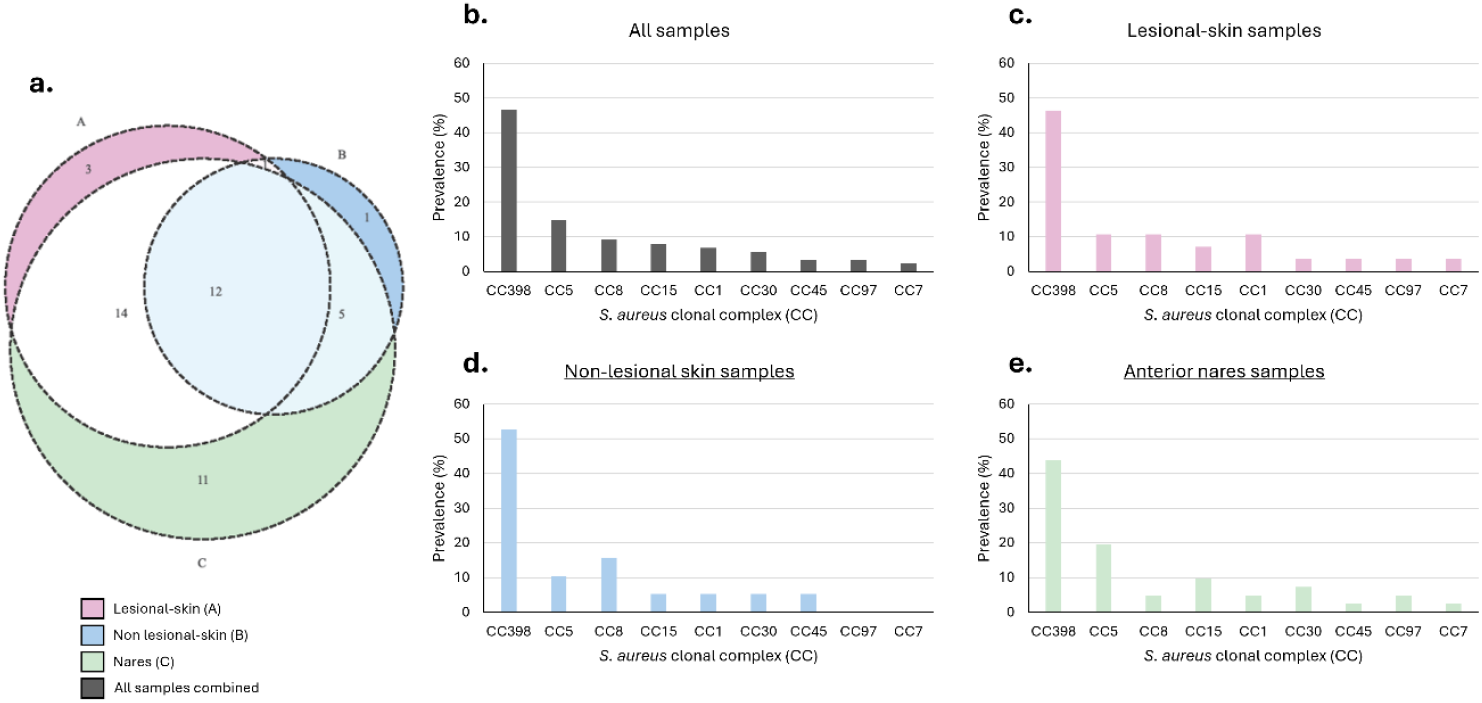
Distribution of *Staphylococcus aureus* colonization sites and prevalence of clonal complexes (CC) in patients with atopic dermatitis (AD). (a) Venn diagram showing the number of patients colonized with *S. aureus* at different body sites: lesional skin (A; pink; n=30), non-lesional skin (B; blue; n=19), and nares (C; green; n=42). Overlapping areas indicate patients colonized at multiple sites. (b–e) Relative frequency of clonal complexes (CCs) among *S. aureus* isolates in (b) all samples combined, (c) lesional skin, (d) non-lesional skin, and (e) nasal samples.

*S. aureus* isolates were characterized by *spa*-typing, from which sequence types (STs) and clonal complexes (CCs) were inferred, and classified as methicillin-susceptible (MSSA) or -resistant (MRSA) through polymerase chain reaction (PCR) detection of *mecA*/*mecC* genes (Supplementary Methods; Supplementary Table S1). *S. aureus* isolates were predominantly MSSA (98%, n = 89/91), with only two MRSA identified (nasal samples, CC5). CC398 was identified as the predominant lineage, accounting for nearly half of all *S. aureus* isolates across all sampled sites (Figure 1b) and individual anatomical sites (Figure 1c-e). All CC398 isolates were MSSA belonging to ST398 or ST398-related and carried the immune evasion cluster (IEC), as detected by PCR, which is consistent with a human-adapted lineage. This lineage has been associated with colonization in vulnerable populations in community settings (Conceição et al. 2019) and invasive and severe infections in Portugal and globally (Gregorio et al. 2024; Nobre et al. 2024), emphasizing its potential clinical relevance. Although MSSA-ST398 has been reported in patients with AD in Spain, Italy, and Canada, its prevalence in our sample was higher than previously described (47% vs. 2.5-33%) (Benito et al. 2015; Sivori et al. 2024; Yeung et al. 2011). The unusually high prevalence of MSSA-ST398 observed in our study suggests a distinctive local epidemiological pattern and raises the possibility that, beyond geographic occurrence, this lineage may be adapted to the AD skin microenvironment. To explore potential factors underlying the success of MSSA-ST398 in AD, we compared its antibiotic resistance profile and virulence potential with those of other CCs identified in our collection (Supplementary Methods; Supplementary Table S1).

Antibiotic susceptibility testing revealed that resistance was most common to penicillin (52%, 47/91) and erythromycin (40%, 36/91), the latter largely driven by MSSA-ST398, accounting for 83% (30/36) of resistant isolates. Distinct pathogenic profiles were observed among *S. aureus* lineages (Figure 2), with ST398 isolates being characterized by reduced biofilm formation (*p* = 0.003; Figure 2a), but strong hemolytic activity (~80%; Figure 2b) and frequent moderate-to-high urease activity (>78%; Figure 2c).

**Figure 2.**
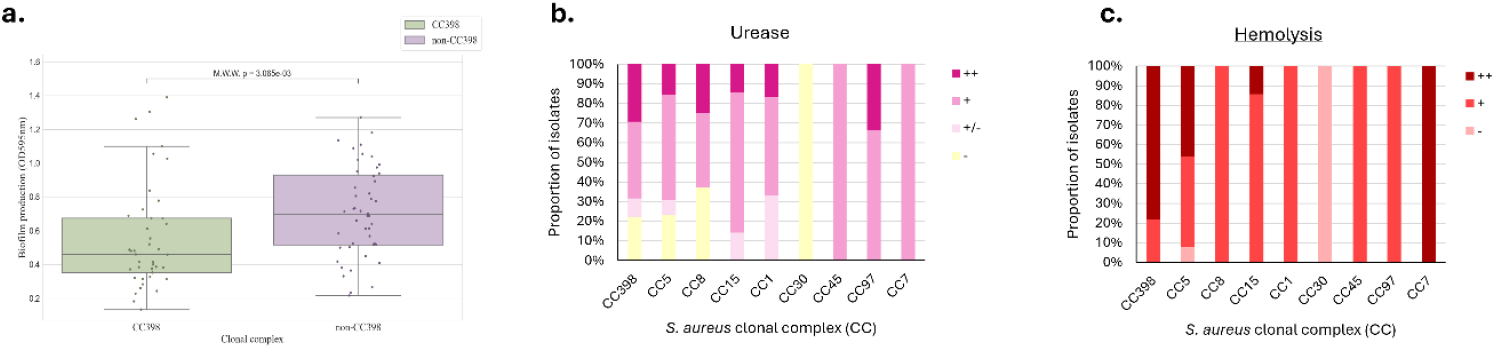
Virulence traits of *Staphylococcus aureus* isolates collected from atopic dermatitis (AD) patients. (a) Biofilm production by *S. aureus* isolates, expressed as optical density at 595 nm, comparing CC398 and non-CC398 lineages. Group differences were assessed using the Mann–Whitney U test, with the *p*-value shown. (b) Urease production across different clonal complexes, classified as non-producer (–), weak (+/–), moderate (+), or strong (++), based on the degree of color change from yellow to pink on Christensen’s urea agar. (c) Hemolysin production across clonal complexes, assessed on sheep blood agar and classified as no hemolysis (–, halo 0 mm), moderate production (+; halo 10–14 mm), or strong production (++; halo ≥15 mm).

This combination of low biofilm formation and high expression of skin barrier– damaging enzymes may confer a selective advantage to MSSA-ST398 in AD skin. Reduced biofilm formation could facilitate bacterial dispersal and promote a more active and immunogenic metabolism, while hemolysin production, known to damage keratinocytes, likely enhances colonization, persistence, and contribution to AD flares. In addition, urease-mediated urea degradation, previously shown to increase skin pH and impair barrier function (Costa and Horswill 2025), may represent a unrecognized virulence mechanism contributing to AD pathogenesis. Together, these findings provide a possible explanation for the success of MSSA-ST398 in AD beyond its local prevalence, while clones lacking AD-adaptive traits may be less competitive.

*S. aureus* colonization in AD reflects an interplay between local molecular epidemiology and disease-specific selective pressures. In Portugal, this has favoured the dominance of the human-adapted MSSA-ST398 lineage, likely driven by virulence traits promoting persistent colonization and skin barrier disruption. Integrating local epidemiology with clonal background and pathogenic traits is therefore critical to understanding *S. aureus*– AD interaction and may inform future strategies to limit bacterial burden and disease exacerbation. Given that samples were derived from a single hospital in Portugal, multicenter and multinational studies will be needed to determine the broader applicability of these findings.

## Ethics Statement

This study was performed in accordance with the Declaration of Helsinki. This human study was approved by Ethics Committee of the University Hospital of Coimbra - approval: Proc 2024-ESI.SF-110. All adult participants provided written informed consent to participate in this study. Samples were collected using non-invasive procedures causing no pain or discomfort to patients. Clinical data collected were anonymized.

## Data Availability Statement

All data collected under the scope of this study are available as part of the core text of this paper or as part of supplemental material. S. aureus whole genome sequence data will be deposited in Short Reads Archive (SRA, reference to be provided soon).

## Acknowledgements

This work was supported by FCT – Fundação para a Ciência e a Tecnologia, I.P., through MOSTMICRO-ITQB R&D Unit (doi.org/10.54499/UID/04612/2025, UID/PRR/4612/2025) and LS4FUTURE Associated Laboratory (DOI 10.54499/LA/P/0087/2020).This study was funded by project ref. 16115 COMPETE2030-FEDER-00726600, LISBOA2030-FEDER-00726600 from Fundação para a Ciência e a Tecnologia and PT2030 regional funds; AB and MA were supported by the PhD Studentships 2022.11966.BD and 2022.14215.BD, respectively, from Fundação para a Ciência e a Tecnologia.

## Author Contributions Statement (CRediT-compliant)

Diana Caieiro (DC) did all the experimental work, did the analysis of data and wrote the manuscript; Nuno Faria (NF) helped in study design, did the analysis of data and reviewed the manuscript; Ana Botelho (AB) did analysis of data and reviewed the manuscript; Mariana Araújo did analysis of data and reviewed the manuscript; Leonor Ramos (LR) collected the samples and clinical data and reviewed the manuscript; Joana Calvão (JC) collected samples and clinical data and reviewed the manuscript; Margarida Gonçalo (MG) did the study design, provided resources, collected the samples and clinical data, discussed the results and reviewed the manuscript; Maria Miragaia (MM) did the study design, provided resources, analyzed and discussed data, supervised experimental work and reviewed the manuscript.

