## Supplementary Material for "Methicillin-Susceptible *Staphylococcus aureus* ST398 in atopic dermatitis in Portugal displays pathogenic traits associated with impaired skin barrier function"

**The methicillin-susceptible *Staphylococcus aureus* ST398 (MSSA-ST398), highly frequent in atopic dermatitis in Portugal, displays unique pathogenic traits**

Diana Caieiro (1), Nuno A. Faria (1), Ana Botelho (1), Mariana Araújo (1), Leonor Ramos (2,3), Joana Calvão (2), Margarida Gonçalo (2,3), Maria Miragaia (1)

(1) Laboratory of Bacterial Evolution and Molecular Epidemiology, Instituto de Tecnologia Química e Biológica António Xavier, Universidade Nova de Lisboa, Oeiras, 2780-157, Portugal

(2) Department of Dermatology, University Hospital, Unidade Local de Saúde de Coimbra, Coimbra, 3000-075, Portugal

(3) Dermatology, Faculty of Medicine, University of Coimbra, Coimbra, 3004-561, Portugal

### SUPPLEMENTARY METHODS

#### Sampling and *S. aureus* isolation procedure

Sample collection was conducted from April 2024 to April 2025 at the Department of Dermatology of the Hospital of Coimbra Local Health Unit, in adult AD patients during their regular visits to the AD specialized consultation. Microbiome samples were collected by dermatologists using a sterile cotton swab (Copan, Brescia, Italy) pre-wettened with 0.85% sterile saline serum from a defined 2 $cm^2$ area of an AD lesion, mostly (79%, n=45/57) on the antecubital flexural area (A), a defined 2 $cm^2$ area of non-lesional skin, mostly (96%, n=55/57) anterior abdominal skin (B) and from one anterior nare (C). No topical therapy or emollients were applied to the sampling areas in the previous 24h. Each set of three swabs collected from each patient was given a code number and sent to the laboratory in Stuart transport medium (Copan) with transportation taking approximately one working day. All samples were processed within one week of their arrival. From each patient, personal data was anonymized, and demographic data, disease duration, AD severity measured by the Eczema Area and Severity Index (EASI), itch intensity in the previous week measured by Worst Itch Numerical Rating Scale (WI-NRS) and current and previous treatment were registered.

After arrival at the laboratory, swabs were resuspended in 1.5 mL of Phosphate Buffered Saline (PBS) 1X. A 50 µL volume of each sample suspension was inoculated onto CHROMagar Staph aureus (CHROMagar, Saint-Denis, France) using glass beads and incubated overnight at 37ºC. This selective chromogenic culture medium allows the presumptive identification of *S. aureus* by the growth of colonies with a mauve coloration. A single colony was selected to proceed the isolation process; if multiple variants of mauve colonies were observed, then the predominant colony morphology was chosen. The selected colony was inoculated onto Mannitol Salt Agar

(MSA) (BD, New Jersey, USA) and incubated overnight at 37 °C. A single presumptive *S. aureus* colony was subsequently cultured on Tryptic Soy Agar (TSA) (BD) and incubated overnight at 37 °C. Staphaurex Latex Agglutination Test (Thermo Fisher Scientific, Massachusetts, USA), which detects clumping factor and protein A positive staphylococci, was performed for all isolates. Confirmed *S. aureus* isolates were stored in TSB with 15% glycerol at –80 °C. Isolates that did not ferment mannitol on MSA were classified as *S. aureus* if they tested positive in the Staphaurex assay and exhibited the presence of the *spa* gene, determined by PCR as described in the next section.

**Fast DNA extraction method**

*S. aureus* DNA was isolated using a fast extraction method. Briefly, three to four colonies were collected from overnight cultures and resuspended in Tris-EDTA (TE) buffer. Lysis was performed using lysostaphin (0.2 mg/mL) followed by incubation at 37°C for 30 minutes. The lysate was subjected to 95°C for 15 minutes to complete lysis and centrifuged at 13000 rpm (5452 Minispin, Eppendorf) for 5 minutes. The supernatant containing genomic DNA was collected and stored at -20°C until further use.

**PCR detection of *S. aureus* protein A (Spa) gene, immune evasion cluster (IEC) and methicillin resistance genes (*mecA*, *mecC*)**

PCR assays were performed to detect genes related to the IEC, methicillin resistance (*mecA*, *mecC*), and the *spa* gene for molecular typing of *S. aureus* isolates (Aires-De-Sousa et al. 2006; Shopsin et al. 1999). All amplifications were conducted in a MiniAmp™ Plus Thermal Cycler (Thermo Fisher Scientific) using a final reaction volume of 50 µL. The PCR reaction was the same for all targets and consisted of 0.025 U/µL GoTaq DNA polymerase (Promega, Wisconsin, USA), 1× PCR buffer, 25 mM $MgCl_2$, 2 mM dNTP mix, and 20 pmol/µL of each primer. PCR products were resolved by electrophoresis on a 1% agarose gel (NZYtech, Lisboa, Portugal) in Tris Acetate EDTA (TAE) 1X supplemented with 0.003% v/v of GreenSafe (NZYTech) and run at 7 V/cm for 30 minutes. DNA was visualized under ultraviolet (UV) Gel-Doc TM EZ Imager (Bio-Rad, California, USA) and images recorded. The molecular weight ladder GeneRuler 1 kb Plus DNA Ladder (Thermo Fisher Scientific) was used.

***S. aureus* protein A genotyping**

The polymorphic X region of *spa* gene of all *S. aureus* isolates was amplified by PCR according to the published protocol, with the annealing temperature modified to 55 °C (Aires-De-Sousa et al. 2006; Shopsin et al. 1999). Primers spa-1113F (TAA AGA CGA TCC TCC GGT TAG G) and spa-1514R (CAG CAG TAG TGC GGT TTG CTT) were used, generating fragments of 110–422 base pairs.

PCR products were purified using the PCRquick-spinTM Kit (iNtRON Biotechnology, Washington, USA) and forward and reverse strands sequenced by Sanger methodology. The *spa* types were assigned by analysing forward and reverse sequence chromatograms in the Ridom Staph Type software as previously described (Harmsen et al. 2003). After determining the *spa* type, the most likely ST was inferred using the following criteria: (1) direct prediction by the software; (2) if unavailable, verification in the Ridom SpaServer database (spaServer.ridom.de) to check for a previously reported ST association; (3) if absent, literature search to identify prior descriptions; (4) if not previously described, assignment to an ST-related type when the *spa* sequence differed by no more than one repeat from a *spa*-type already linked to that ST. If none of these criteria were met, or if multiple STs were proposed by the software, the ST was designated as unknown. The most likely clonal complex (CC) was inferred based on the predictions from RidomStaph and literature.

**IEC typing of MSSA-ST398 isolates**

To assess the presence of the IEC and determine the likely origin (human or animal) of all MSSA-ST398 isolates, PCR assays targeting *Sa3int*, *chp*, *scn*, *sak*, *sea*, and *sep* genes were performed. For *Sa3int*, *chp*, *scn*, and *sak*, PCR was performed according to Van Wamel, Goerke and co-workers (Goerke et al. 2009; Van Wamel et al. 2006) using the following thermal cycling parameters: initial denaturation (4 minutes at 94°C), followed by 30 cycles of denaturation (30 seconds at 94°C), annealing (30 seconds at 50°C), and extension (1 minute at 72°C), and finishing with a single extension (10 minutes at 72°C). For amplification of the *sea* gene, the annealing was adjusted to 1 minute at 60 °C, while for *sep* it was set to 1 minute at 50 °C. For detection of the *scn*, *chp*, *sak*, and *sep* genes, PCR assays were performed using the primer pairs Scn-1/Scn-2, Chp-1/Chp-2, Sak-1/Sak-2, and Gsep-1/Gsep-2, generating fragments of 258, 366, 223, and 182 bp, respectively, with *S. aureus* strain N315 (Van Wamel et al. 2006) used as the control strain for all assays. Detection of the *sea* gene was carried out using primers SEA F and SEA R, producing a 520 bp fragment, with *S. aureus* strain FRI913 (Monday and Bohach 1999) used as the control.

***mecA* and *mecC* search**

To classify *S. aureus* isolates as MSSA or MRSA, a *mecA* and *mecC* amplification by PCR was performed, using the following thermal cycling conditions: initial denaturation (4 min at 94ºC), followed by 30 cycles of denaturation (30 seconds at 94ºC), annealing (1 minute at 57ºC for *mecA* and 52ºC for *mecC*), and extension (90 seconds at 72ºC), and finishing with a single extension (10 minutes at 72ºC). For detection of the *mecA* gene, primers mecA1 (GGT CCC ATT AAC TCT GAA G) and mecA2 (AGT TCT CGA GTA CCG GAT TTG) were used, producing a 1039 bp fragment, with *S. aureus* strain MW2 as the control(Miragaia, M, Couto, I, De Lencastre 2005).

For detection of the *mecC* gene, primers mecC F3 (ACA CCT TTT AGG TTA TGT GG) and mecC R8 (AAC CAA CCT ATT TGT CTT CC) were used, generating a 1101 bp fragment, with strain LGA251 (García-álvarez et al. 2003) as the control. Isolates were considered MRSA if either gene was detected, and MSSA if both were absent.

### Antimicrobial susceptibility testing

All *S. aureus* isolates were tested for antimicrobial susceptibility using disk diffusion method (Kirby-Bauer), as described in the European Committee on Antimicrobial Susceptibility Testing (EUCAST) guidelines (v14.0, January 2024). The panel of 19 antibiotics (OXOID, Basingstoke, UK) included: benzylpenicillin (BEN, 10 Units), ceftaroline (CTL, 5 µg), cefoxitin (CXI, 30 µg), ciprofloxacin (CIP, 5 µg), clindamycin (CLI, 2 µg), erythromycin (ERY, 15 µg), fusidic acid (FUS, 10 µg), gentamicin (GEN, 10 µg), kanamycin (KAN, 30 µg), linezolid (LIN, 10 µg), minocycline (MIN, 30 µg), mupirocin (MUP, 200 µg), norfloxacin (NOR, 10 µg), oxacillin (OXA, 1 µg), rifampicin (RIF, 5 µg), tetracycline (TET, 15 µg), trimethoprim-sulfamethoxazole (TRS, 25 µg), tigecycline (TIG, 15 µg) and vancomycin (VAN, 30 µg). The breakpoints were those described in EUCAST (January 2024) except for oxacillin, vancomycin (Clinical & Laboratory Standards Institute, CLSI, 2008) and mupirocin (British Society for Antimicrobial Chemotherapy, BSAC, 2011).

### Static biofilm assay

Biofilm formation ability of all *S. aureus* isolates was evaluated as previously described (Kwasny and Opperman 2010) using the microtiter assay (Corning, Corning, USA). Briefly, overnight cultures were diluted 1:100 with TSB (BD) supplemented with 1% glucose. After incubation in static conditions at 37 ºC for 18 hours, the wells were washed with deionized water to remove planktonic growth and nonadherent bacteria, and the biofilms were heat fixed at 60 ºC for 60 minutes. The fixed biofilms were stained with 0.06% crystal violet. After 5 minutes of staining reaction, a 30% acetic acid solution was added to each well and the optical density was measured at 595 nm in the microtiter plate reader InfiniteR 200 PRO series (Tecan Group Ltd.). *S. epidermidis* strains ATCC 35984 and 19N (Gonçalves et al. 2022) were used as positive and negative control, respectively, for biofilm production ability. For each isolate three technical replicas and three biological replicas were performed.

### Urease production assay

To test for urease production, a heavy inoculum (10 µL loop) of all isolates was streaked in Christensen's Urea Agar (Sigma-Aldrich) supplemented with urea (Merck), according to manufacturer instructions. Plates were incubated at 37ºC for 24 hours. Medium plates without urea were used as a control. The test was considered negative when there was no change in medium colour, suggesting that the bacteria did not metabolize urea. Based on the degree of colour change from yellow to pink, the isolates were categorized into groups reflecting different levels of urease activity: no producer (no change of colour, same colour as medium control), weak producer (slight change of colour, most often only in a small area of the plate), moderate producer (all or almost all area of the plate has undergone colour change) or strong producer (the entire plate changed colour to a very strong tone).

**Hemolysin production assay**

To test for hemolytic activity of all *S. aureus* isolates, bacterial colonies grown overnight in TSA (BD) were resuspended in a 0.85% saline solution to obtain a 0.5 McFarland standard inoculum suspension. A drop of 10 µL was spotted onto sheep blood agar (BioMérieux, Marcy-l'Étoile, France) and cultures were incubated at 37 ºC for 24 hours. A negative result, or gamma-hemolysis, corresponded to no change in the appearance of the agar around the bacterial spot. A positive result corresponded to a clear and colourless zone around the spot (complete lysis of red blood cells, beta-hemolysis) or to a greenish/brownish discoloration in the agar (partial lysis of red blood cells, alpha-hemolysis). *Streptococcus pneumoniae* strain ATCC49619 and *Streptococcus pyogenes* strain ATCC12344 were used as a control for alpha and beta hemolysis, respectively. To quantify the extent of hemolysis, the halo zone was measured and recorded. Three groups of hemolysis were established, depending on the size of the halo produced: gamma or no hemolysis if no halo was present (-); moderate producer if the halo size was between 10 and 14 mm (+); strong producer if the halo size was higher than 15 mm (++).

**Whole Genome Sequencing**

Bacterial genomic DNA was extracted from eight MSSA-ST398 isolates obtained from four patients, including paired skin lesion and anterior nares isolates from each patient, using the DNeasy Blood & Tissue Kit (Qiagen, Hilden, Germany) according to the manufacturer's instructions. Whole-genome sequencing was performed on an Illumina NextSeq 2000 platform (Illumina, CA, USA) using the NextSeq P1 300-cycle kit with paired-end chemistry (2 × 150 bp), targeting an average coverage depth of approximately 100X. Sequencing libraries were prepared using the Nextera XT DNA Library Preparation Kit (Illumina). Raw Illumina reads were grouped into forward and reverse reads, quality-checked, and *de novo* assembled using the INNUca pipeline (version 4.2.2) with default parameters (https://github.com/B-UMMI/INNUca). The real coverage of *de novo* assemblies varied between 71 and 193, and each draft genome had between 28 and 54 contigs.

**Phylogenetic analysis**

The ST of the 19 isolates selected for WGS was further confirmed both by INNUca and by uploading contigs to the Pathogenwatch watch platform (version 23.5.0) available at https://pathogen.watch/. Phylogenetic relatedness among *S. aureus* isolates (n = 8) was inferred based on concatenated alignments of core single-nucleotide polymorphisms (SNPs) using CSIPhylogeny (Kaas et al. 2014). The following parameters were applied: minimum depth at SNP positions, 10×; minimum relative depth, 10%; minimum SNP quality, 30; minimum read mapping quality, 25; minimum Z-score, 1.96; and minimum distance between SNPs, disabled. One isolate from the collection (2A) was used as the reference genome. A total of 2.652.939 positions were found in all analysed genomes, corresponding to 97.5% coverage of the reference genome by all isolates.

### SUPPLEMENTARY TABLES

**Supplementary Table S1** Molecular characteristics, antimicrobial resistance, and virulence-associated phenotypes of *Staphylococcus aureus* isolates recovered from atopic dermatitis (AD) patients.

| Sample ID | *spa*-type | ST | CC | Antibiotic resistance | Biofilm OD | Urease | Hemolysis halo | Hemolysis group | *mecA* | *mecC* | MRSA/MSSA |
|---|---|---|---|---|---|---|---|---|---|---|---|
| **1A** | t279 | ST15 | CC15 | NOR, BEN | 0.518 | + | 10 | + | no | no | MSSA |
| **1C** | t224 | ST97 | CC97 | BEN | 0.788 | + | 14 | + | no | no | MSSA |
| **2A** | t1451 | ST398 | CC398 | ERY | 0.400 | + | 17 | ++ | no | no | MSSA |
| **2B** | t1451 | ST398 | CC398 | ERY, CLI* | 0.555 | + | 17 | ++ | no | no | MSSA |
| **2C** | t1451 | ST398 | CC398 | ERY, CLI* | 0.259 | + | 17 | ++ | no | no | MSSA |
| **3A** | t400 | ST8 | CC8 | BEN | 0.732 | ++ | 11 | + | no | no | MSSA |
| **3B** | t400 | ST8 | CC8 | FUS, BEN | 0.687 | ++ | 11 | + | no | no | MSSA |
| **4A** | t840 | ST30, ST378 | CC30 | BEN | 0.820 | - | 0 | - | no | no | MSSA |
| **4B** | t840 | ST30, ST378 | CC30 | BEN | 0.523 | - | 0 | - | no | no | MSSA |
| **4C** | t840 | ST30, ST378 | CC30 | BEN | 0.716 | - | 0 | - | no | no | MSSA |
| **6C** | t021 | ST30, ST33, ST55 | CC30 | BEN | 0.640 | - | 0 | - | no | no | MSSA |
| **7A** | t535 | ST5 | CC5 | BEN | 0.614 | + | 15 | ++ | no | no | MSSA |
| **7C** | t535 | ST5 | CC5 | BEN | 0.409 | + | 15 | ++ | no | no | MSSA |
| **8A** | t1451 | ST398 | CC398 | ERY | 0.320 | + | 15 | ++ | no | no | MSSA |
| **8B** | t1451 | ST398 | CC398 | ERY, CLI* | 0.314 | + | 15 | ++ | no | no | MSSA |
| **8C** | t1451 | ST398 | CC398 | ERY, CLI* | 0.481 | + | 15 | ++ | no | no | MSSA |
| **9A** | t6587 | ST398 | CC398 | - | 0.384 | - | 13 | + | no | no | MSSA |
| **9C** | t6587 | ST398 | CC398 | - | 0.415 | - | 13 | + | no | no | MSSA |

For each isolate, the table reports: sample ID; *spa* type; inferred multilocus sequence type (ST); clonal complex (CC); antibiotic resistance profile; biofilm production (optical density, OD); urease production; hemolysis halo diameter (mm); hemolysis classification; presence of *mecA* and *mecC* genes; and classification as methicillin-resistant (MRSA) or methicillin-susceptible (MSSA) *S. aureus*. Sampling sites are indicated by the sample ID suffix: A, AD lesion; B, non-lesional skin; C, nares. Antibiotic resistance abbreviations: BEN: Benzylpenicillin/penicillin G, ERY: Erythromycin, CLI: Clindamycin, FUS: Fusidic acid, KAN: Kanamycin, MUP: Mupirocin, TET: Tetracycline, NOR: Norfloxacin, CXI: Cefoxitin, RIF: Rifampicin, MIN: Minocycline, CIP: Ciprofloxacin, GEN: Gentamycin. An asterisk (*) indicates inducible resistance (D-test positive). Urease production levels are indicated as follows: –, no production; –/+, weak production; +, moderate production; ++, strong production. Hemolysis halo values represent the diameter of the clear zone surrounding colonies on blood agar. Hemolysin production levels are indicated as follows: –, no production; + moderate producer; ++ strong producer.

**Supplementary Table S1 (cont.)** Molecular characteristics, antimicrobial resistance, and virulence-associated phenotypes of *Staphylococcus aureus* isolates recovered from atopic dermatitis (AD) patients.

| Sample ID | *spa*-type | ST | CC | Antibiotic resistance | Biofilm OD | Urease | Hemolysis halo | Hemolysis group | *mecA* | *mecC* | MRSA/MSSA |
|---|---|---|---|---|---|---|---|---|---|---|---|
| **12C** | t571 | ST398 | CC398 | ERY, CLI* | 0.673 | - | 12 | + | no | no | MSSA |
| **13A** | t177 | ST3 | CC1 | FUS | 1.039 | +/- | 12 | + | no | no | MSSA |
| **13C** | t177 | ST3 | CC1 | FUS, BEN | 0.805 | +/- | 12 | + | no | no | MSSA |
| **14C** | t1451 | ST398 | CC398 | RIF, BEN | 0.283 | + | 15 | ++ | no | no | MSSA |
| **15C** | t688 | ST5 | CC5 | CXI, MIN, FUS, TET, ERY, CLI, BEN | 0.990 | + | 15 | ++ | yes | no | MRSA |
| **16C** | t346 | ST15 | CC15 | BEN | 0.855 | + | 11 | + | no | no | MSSA |
| **17A** | t13998 | ST72 | CC8 | MUP, BEN | 0.449 | - | 14 | + | no | no | MSSA |
| **17B** | new spa-type | ST72-related | CC8 | MUP, BEN | 0.567 | - | 14 | + | no | no | MSSA |
| **17C** | t13998 | ST72 | CC8 | BEN | 0.523 | - | 14 | + | no | no | MSSA |
| **18A** | t9061 | unknown | unknown | BEN | 0.347 | ++ | 11 | + | no | no | MSSA |
| **23A** | t267 | ST97 | CC97 | BEN | 0.612 | + | 14 | + | no | no | MSSA |
| **23C** | t267 | ST97 | CC97 | BEN | 0.713 | ++ | 14 | + | no | no | MSSA |
| **24A** | t430 | ST8 | CC8 | FUS | 0.951 | + | 13 | + | no | no | MSSA |
| **24C** | t430 | ST8 | CC8 | FUS, PEN | 0.730 | + | 11 | + | no | no | MSSA |
| **25B** | t2041 | ST8 | CC8 | FUS | 0.364 | + | 12 | + | no | no | MSSA |
| **25C** | t002 | ST5, ST231 | CC5 | - | 1.272 | + | 15 | ++ | no | no | MSSA |
| **26A** | t084 | ST15, ST18 | CC15 | KAN, TET, ERY, CLI*, BEN | 0.499 | + | 12 | + | no | no | MSSA |
| **26B** | t084 | ST15, ST18 | CC15 | KAN, TET, ERY, CLI*, BEN | 0.418 | + | 12 | + | no | no | MSSA |
| **26C** | t084 | ST15, ST18 | CC15 | KAN, TET, ERY, CLI*, BEN | 0.502 | + | 12 | + | no | no | MSSA |
| **27A** | t1451 | ST398 | CC398 | ERY | 0.314 | ++ | 11 | + | no | no | MSSA |

For each isolate, the table reports: sample ID; *spa* type; inferred multilocus sequence type (ST); clonal complex (CC); antibiotic resistance profile; biofilm production (optical density, OD); urease production; hemolysis halo diameter (mm); hemolysis classification; presence of *mecA* and *mecC* genes; and classification as methicillin-resistant (MRSA) or methicillin-susceptible (MSSA) *S. aureus*. Sampling sites are indicated by the sample ID suffix: A, AD lesion; B, non-lesional skin; C, nares. Antibiotic resistance abbreviations: BEN: Benzylpenicillin/penicillin G, ERY: Erythromycin, CLI: Clindamycin, FUS: Fusidic acid, KAN: Kanamycin, MUP: Mupirocin, TET: Tetracycline, NOR: Norfloxacin, CXI: Cefoxitin, RIF: Rifampicin, MIN: Minocycline, CIP: Ciprofloxacin, GEN: Gentamycin. An asterisk (*) indicates inducible resistance (D-test positive). Urease production levels are indicated as follows: –, no production; –/+, weak production; +, moderate production; ++, strong production. Hemolysis halo values represent the diameter of the clear zone surrounding colonies on blood agar. Hemolysin production levels are indicated as follows: –, no production; + moderate producer; ++ strong producer.

**<u>Supplementary Table S1 (cont.)</u>** Molecular characteristics, antimicrobial resistance, and virulence-associated phenotypes of *Staphylococcus aureus* isolates recovered from atopic dermatitis (AD) patients.

| Sample ID | *spa*-type | ST | CC | Antibiotic resistance | Biofilm OD | Urease | Hemolysis halo | Hemolysis group | *mecA* | *mecC* | MRSA/MSSA |
|---|---|---|---|---|---|---|---|---|---|---|---|
| **27B** | t1451 | ST398 | CC398 | ERY | 0.243 | ++ | 11 | + | no | no | MSSA |
| **27C** | t1451 | ST398 | CC398 | ERY | 0.182 | ++ | 11 | + | no | no | MSSA |
| **29A** | t015 | ST45 | CC45 | BEN | 0.233 | + | 10 | + | no | no | MSSA |
| **29C** | t002 | ST5, ST231 | CC5 | CXI, FUS, NOR, CIP, ERY, CLI* | 0.700 | +/- | 11 | + | yes | no | MRSA |
| **30B** | t015 | ST45 | CC45 | MUP, BEN | 0.331 | + | 11 | + | no | no | MSSA |
| **32A** | t1451 | ST398 | CC398 | KAN, GEN, RIF, ERY, BEN | 0.326 | ++ | 18 | ++ | no | no | MSSA |
| **32B** | t1451 | ST398 | CC398 | KAN, GEN, ERY | 0.352 | ++ | 18 | ++ | no | no | MSSA |
| **32C** | t1451 | ST398 | CC398 | KAN, GEN, ERY, BEN | 0.520 | ++ | 18 | ++ | no | no | MSSA |
| **33A** | t571 | ST398 | CC398 | ERY, CLI* | 0.133 | + | 17 | ++ | no | no | MSSA |
| **33B** | t571 | ST398 | CC398 | ERY | 0.228 | + | 17 | ++ | no | no | MSSA |
| **33C** | t230 | ST45 | CC45 | - | 0.266 | + | 11 | + | no | no | MSSA |
| **34A** | t15167 | ST398-related | CC398 | ERY | 0.725 | + | 16 | ++ | no | no | MSSA |
| **35C** | t803 | ST15 | CC15 | FUS | 0.583 | +/- | 16 | ++ | no | no | MSSA |
| **36B** | t1451 | ST398 | CC398 | FUS, ERY | 0.480 | +/- | 18 | ++ | no | no | MSSA |
| **36C** | t1451 | ST398 | CC398 | ERY | 0.492 | + | 18 | ++ | no | no | MSSA |
| **37A** | t571 | ST398 | CC398 | MUP | 0.371 | + | 17 | ++ | no | no | MSSA |
| **37B** | t571 | ST398 | CC398 | - | 0.612 | + | 17 | ++ | no | no | MSSA |
| **37C** | t002 | ST5, ST231 | CC5 | - | 1.109 | + | 17 | ++ | no | no | MSSA |
| **38C** | t793 | unknown | CC15 | BEN | 0.775 | ++ | 11 | + | no | no | MSSA |
| **40A** | t571 | ST398 | CC398 | ERY, CLI* | 1.026 | - | 15 | ++ | no | no | MSSA |

For each isolate, the table reports: sample ID; *spa* type; inferred multilocus sequence type (ST); clonal complex (CC); antibiotic resistance profile; biofilm production (optical density, OD); urease production; hemolysis halo diameter (mm); hemolysis classification; presence of *mecA* and *mecC* genes; and classification as methicillin-resistant (MRSA) or methicillin-susceptible (MSSA) *S. aureus*. Sampling sites are indicated by the sample ID suffix: A, AD lesion; B, non-lesional skin; C, nares. Antibiotic resistance abbreviations: BEN: Benzylpenicillin/penicillin G, ERY: Erythromycin, CLI: Clindamycin, FUS: Fusidic acid, KAN: Kanamycin, MUP: Mupirocin, TET: Tetracycline, NOR: Norfloxacin, CXI: Cefoxitin, RIF: Rifampicin, MIN: Minocycline, CIP: Ciprofloxacin, GEN: Gentamycin. An asterisk (*) indicates inducible resistance (D-test positive). Urease production levels are indicated as follows: –, no production; –/+, weak production; +, moderate production; ++, strong production. Hemolysis halo values represent the diameter of the clear zone surrounding colonies on blood agar. Hemolysin production levels are indicated as follows: –, no production; + moderate producer; ++ strong producer.

**<u>Supplementary Table S1 (cont.)</u>** Molecular characteristics, antimicrobial resistance, and virulence-associated phenotypes of *Staphylococcus aureus* isolates recovered from atopic dermatitis (AD) patients.

| Sample ID | *spa*-type | ST | CC | Antibiotic resistance | Biofilm OD | Urease | Hemolysis halo | Hemolysis group | *mecA* | *mecC* | MRSA/MSSA |
|---|---|---|---|---|---|---|---|---|---|---|---|
| **40B** | t571 | ST398 | CC398 | ERY, CLI* | 0.778 | + | 19 | ++ | no | no | MSSA |
| **40C** | t571 | ST398 | CC398 | ERY, CLI* | 1.055 | +/- | 16 | ++ | no | no | MSSA |
| **42C** | t571 | ST398 | CC398 | NOR | 0.640 | ++ | 16 | ++ | no | no | MSSA |
| **44A** | t002 | ST5, ST231 | CC5 | BEN | 1.181 | - | 13 | + | no | no | MSSA |
| **44B** | t002 | ST5, ST231 | CC5 | BEN | 1.135 | - | 13 | + | no | no | MSSA |
| **44C** | t002 | ST5, ST231 | CC5 | BEN | 1.088 | - | 13 | + | no | no | MSSA |
| **45A** | t1451 | ST398 | CC398 | PEN, ERY | 1.261 | ++ | 16 | ++ | no | no | MSSA |
| **45C** | t1451 | ST398 | CC398 | PEN, ERY | 1.305 | ++ | 15 | ++ | no | no | MSSA |
| **46A** | t3092 | unknown | unknown | FUS | 1.041 | ++ | 15 | ++ | no | no | MSSA |
| **47B** | t002 | ST5, ST231 | CC5 | BEN | 0.380 | ++ | 11 | + | no | no | MSSA |
| **47C** | t18827 | unknown | unknown | FUS, BEN | 1.096 | - | 13 | + | no | no | MSSA |
| **48A** | t571 | ST398 | CC398 | - | 1.389 | - | 17 | ++ | no | no | MSSA |
| **48C** | t571 | ST398 | CC398 | - | 1.100 | - | 17 | ++ | no | no | MSSA |
| **51C** | t306 | ST5 | CC5 | - | 0.893 | + | 11 | + | no | no | MSSA |
| **52A** | t127 | ST1 | CC1 | FUS, BEN | 1.020 | + | 11 | + | no | no | MSSA |
| **52B** | t127 | ST1 | CC1 | FUS, BEN | 1.054 | + | 11 | + | no | no | MSSA |
| **52C** | t127 | ST1 | CC1 | FUS, BEN | 0.940 | + | 11 | + | no | no | MSSA |
| **55C** | t567 | ST398 | CC398 | BEN | 0.674 | ++ | 13 | + | no | no | MSSA |
| **56A** | t5635 | ST398 | CC398 | ERY, CLI* | 0.375 | + | 18 | ++ | no | no | MSSA |
| **56C** | t5635 | ST398 | CC398 | ERY, CLI* | 0.487 | + | 18 | ++ | no | no | MSSA |

For each isolate, the table reports: sample ID; *spa* type; inferred multilocus sequence type (ST); clonal complex (CC); antibiotic resistance profile; biofilm production (optical density, OD); urease production; hemolysis halo diameter (mm); hemolysis classification; presence of *mecA* and *mecC* genes; and classification as methicillin-resistant (MRSA) or methicillin-susceptible (MSSA) *S. aureus*. Sampling sites are indicated by the sample ID suffix: A, AD lesion; B, non-lesional skin; C, nares. Antibiotic resistance abbreviations: BEN: Benzylpenicillin/penicillin G, ERY: Erythromycin, CLI: Clindamycin, FUS: Fusidic acid, KAN: Kanamycin, MUP: Mupirocin, TET: Tetracycline, NOR: Norfloxacin, CXI: Cefoxitin, RIF: Rifampicin, MIN: Minocycline, CIP: Ciprofloxacin, GEN: Gentamycin. An asterisk (*) indicates inducible resistance (D-test positive). Urease production levels are indicated as follows: –, no production; –/+, weak production; +, moderate production; ++, strong production. Hemolysis halo values represent the diameter of the clear zone surrounding colonies on blood agar. Hemolysin production levels are indicated as follows: –, no production; + moderate producer; ++ strong producer.

**Supplementary Table S1 (cont.)** Molecular characteristics, antimicrobial resistance, and virulence-associated phenotypes of *Staphylococcus aureus* isolates recovered from atopic dermatitis (AD) patients.

| Sample ID | *spa*-type | ST | CC | Antibiotic resistance | Biofilm OD | Urease | Hemolysis halo | Hemolysis group | *mecA* | *mecC* | MRSA/MSSA |
|---|---|---|---|---|---|---|---|---|---|---|---|
| **58A** | t091 | ST7 | CC7 | BEN | 0.974 | + | 17 | ++ | no | no | MSSA |
| **58C** | t091 | ST7 | CC7 | BEN | 0.688 | + | 17 | ++ | no | no | MSSA |
| **59B** | t571 | ST398 | CC398 | - | 0.386 | - | 12 | + | no | no | MSSA |
| **59C** | t571 | ST398 | CC398 | - | 0.460 | - | 12 | + | no | no | MSSA |
| **61A** | t571 | ST398 | CC398 | ERY, CLI* | 0.381 | +/- | 16 | ++ | no | no | MSSA |
| **61C** | t571 | ST398 | CC398 | ERY, CLI* | 0.414 | +/- | 16 | ++ | no | no | MSSA |
| **62C** | t1451 | ST398 | CC398 | ERY, CLI* | 0.458 | - | 15 | ++ | no | no | MSSA |
| **63A** | t895 | ST5 | CC5 | BEN | 0.660 | + | 18 | ++ | no | no | MSSA |
| **63C** | t895 | ST5 | CC5 | BEN | 0.216 | ++ | 0 | - | no | no | MSSA |
| **64B** | t11767 | ST398-related | CC398 | KAN, GEN, ERY, CLI*, BEN | 0.688 | ++ | 18 | ++ | no | no | MSSA |
| **64C** | t1451 | ST398 | CC398 | KAN, GEN, ERY, CLI*, BEN | 0.838 | ++ | 18 | ++ | no | no | MSSA |
| **67A** | t2473 | ST1 | CC1 | ERY, CLI* | 0.516 | ++ | 11 | + | no | no | MSSA |
| **67C** | unknown | ST30 | CC30 | BEN | 0.922 | - | 0 | - | no | no | MSSA |

For each isolate, the table reports: sample ID; *spa* type; inferred multilocus sequence type (ST); clonal complex (CC); antibiotic resistance profile; biofilm production (optical density, OD); urease production; hemolysis halo diameter (mm); hemolysis classification; presence of *mecA* and *mecC* genes; and classification as methicillin-resistant (MRSA) or methicillin-susceptible (MSSA) *S. aureus*. Sampling sites are indicated by the sample ID suffix: A, AD lesion; B, non-lesional skin; C, nares. Antibiotic resistance abbreviations: BEN: Benzylpenicillin/penicillin G, ERY: Erythromycin, CLI: Clindamycin, FUS: Fusidic acid, KAN: Kanamycin, MUP: Mupirocin, TET: Tetracycline, NOR: Norfloxacin, CXI: Cefoxitin, RIF: Rifampicin, MIN: Minocycline, CIP: Ciprofloxacin, GEN: Gentamycin. An asterisk (*) indicates inducible resistance (D-test positive). Urease production levels are indicated as follows: –, no production; –/+, weak production; +, moderate production; ++, strong production. Hemolysis halo values represent the diameter of the clear zone surrounding colonies on blood agar. Hemolysin production levels are indicated as follows: –, no production; + moderate producer; ++ strong producer.

**<u>Supplementary Table S2</u>** Pairwise single-nucleotide polymorphism (SNP) distance matrix of methicillin-susceptible *S. aureus* sequence type (ST) 398 (MSSA) isolates collected from the lesional skin (A) and anterior nares (C) of four atopic dermatitis (AD) patients. Distances were calculated using CSIPhylogeny; within-patient comparisons are shown in **bold**.

| | | Nares | | | |
|---|---|---|---|---|---|
| | | **2C** | **8C** | **27C** | **40C** |
| Lesional skin | **2A** | **2** | 162 | 302 | 787 |
| | **8A** | 172 | **14** | 300 | 763 |
| | **27A** | 297 | 285 | **9** | 908 |
| | **40A** | 789 | 763 | 919 | **16** |
